# Age-dependent inequalities in HIV/STI burden and care receipt among men and transgender persons who have sex with men in Nairobi

**DOI:** 10.1101/2021.06.23.21259373

**Authors:** Adrian D Smith, Elizabeth Fearon, Rhoda Kabuti, Erastus Irungu, Mary Kungu, Hellen Babu, Chrispo Nyabuto, Peter Muthoga, Peter Weatherburn, Adam Bourne, Joshua Kimani

## Abstract

**Background:** Gay, bisexual and other men who have sex with men (GBMSM) and transgender persons (TP) bear high burdens of HIV and other sexually transmitted infections (STIs) in sub-Saharan Africa, yet evidence of HIV care coverage for these groups is sparse from the region despite prevailing stigma and discrimination towards these groups.

**Methods:** 618 GBMSM/TP were recruited in Nairobi between May to December 2017 using respondent-driven sampling. Participants reported recent sexual behaviour, HIV testing and care receipt, and symptoms of STIs. Participants tested for HIV using Kenyan testing algorithms and GeneXpert methods, syphilis, viral hepatitis and ano-genital gonorrhoea and chlamydia. We assessed associations with HIV status and detectable HIV viral load using multivariable robust Poisson regression models.

**Findings:** 26.4% (286/618) were HIV positive of whom 76.5% were status aware, 65.3% were on ART, and 47.4% were virally suppressed (<50 copies/ml). Participants 18-22 years old were less likely to be status aware, be receiving ART or to have achieved viral suppression. Mean log viral load was 3.14 log higher in 18-22 year olds compared to older participants. Bacterial STIs were frequently detected at both urethral and rectal sites and a majority of infections at both sites were asymptomatic by self-report (rectal 82.2%, urethral 90.8%).

**Interpretation:** Engagement in the HIV diagnosis and care cascade among GBMSM/TP in Kenya is markedly better than in most sub-Saharan African countries. However it falls short of achievements among the general population in the country and cascades achieved in GBMSM in high income settings. Young men and transgender persons who have sex with men are least well served by the current configuration of adult key population services, and programmes should identify and address the sexual, social and developmental needs of adolescent and young key populations

## Background

Gay, bisexual and other men who have sex with men (GBMSM) and transgender persons (TP) bear disproportionate burdens of HIV risk and HIV infection around the world^1-3^, including in generalised epidemic settings in sub-Saharan Africa ^4,5^. Structural and cultural obstacles, including criminalisation, institutional homophobia and societal antipathy towards these groups continue to challenge efforts to provide equitable access to effective HIV prevention and treatment, particularly in sub-Saharan Africa^6^. International agencies highlight the harmful consequences of unequal access to prevention and treatment upon members of these populations and to efforts to curb national HIV epidemics^7^. Yet despite clear targets for increasing status awareness and anti-retroviral therapy (ART) uptake among key populations ^8^, very few sub-Saharan African countries conduct surveillance to monitor the effectiveness and coverage of treatment programmes for these populations ^9,10^.

Kenya has a declining generalised epidemic with an adult prevalence estimated at 4.9% in 2018, comprehensive national prevention and treatment responses including oral pre-exposure prophylaxis (PrEP), post-exposure prophylaxis (PEP), voluntary male circumcision, test and treat, and broad availability of viral load testing to support HIV care^11^. The Kenya population - based HIV impact Assessment (KENPHIA) study demonstrated the progress toward achievement of UNAIDS 90-90-90 targets in a national survey of Kenya adults ^12^: in 2018, 79.5% of adult PLWHA (15-49 yrs) were aware of their HIV status, of whom 90.6% were receiving ART, of whom 90.9% (or 72% of all PLWH) were virally suppressed ^12^.

Kenya has 15 years of research describing HIV burden among men who have sex with men. In Nairobi, the most recent estimate of HIV prevalence (based on data collected in 2010) was 18.2% – over three times the prevalence amongst the general population ^13^. National AIDS control policies include strategic goals to enhance HIV prevention and treatment service response for GBMSM in line with the WHO recommended package of key population interventions^14,15^. This has enabled a mixed model of prevention and care delivery through non-governmental organisations, private providers and state clinics largely concentrated in major cities. Whilst some providers monitor care retention and outcomes among their clients, the population coverage and quality of HIV care for GBMSM/TP is unknown.

We aimed to estimate the prevalence of HIV and other sexually transmitted infections in a population representative sample of cisgender male and transgender persons who have sex with men living in Nairobi, describe the HIV care cascade and viral load among GBMSM and TP living with HIV, and assess correlations with prevalence of both HIV infection and detectable viraemia in this context.

## Methods

### Recruitment and sampling

Respondent driven sampling was used to recruit 618 participants between May and December 2017 following established methods^16^. Seed participants were referred to the study by three community organisations who provide services to GBMSM communities in Nairobi (GALCK, ISHTAR and HOYMAS). Following formative qualitative research, ten seeds were chosen to optimise diversity in age, marital status, gender identity, socioeconomic status and district of residence within Nairobi County.

Each participant was issued two recruitment coupons and instructions on how to recruit further eligible participants from their social networks. Inclusion criteria were: possession of a valid study coupon; age 18 or over; male gender assignment at birth or current identification as a man; residence within 50km of Nairobi, and consensual anal or oral sexual activity with a man in the previous twelve months. Coupons detailed the location and contact details for the study site but disclosed no information about the purpose of the study. Coupons were uniquely numbered to verify recruiter-recruit links and coupon legitimacy. The opportunity for coupon duplication was reduced by use of non-standard grade watermarked paper, date stamping and limited period of validity after issue. Participants were reimbursed 300 Kenya shillings (∼USD $3) for each recruit they referred to the study who subsequently participated.

### Study procedures

Coupon recipients who satisfied eligibility criteria underwent informed consent procedures with study staff. Recipients were ineligible if they reported coupon receipt from a stranger, coercion to attend or previous participation in the study. Unique identity was established using a commercially available digital fingerprint scanner.

Personal behaviour was collected via self-completed SurveyGizmo™ questionnaire implemented in English and Kiswahili on touch-screen tablets. The questionnaire covered multiple domains including demographic characteristics; sexual behaviour; alcohol and other substance use; knowledge of HIV transmission risks; use of existing HIV/STI prevention methods; recent anogenital symptoms suggestive of STI; experiences of sexuality-related stigma, discrimination or violence^17,18^. In addition, the questionnaire included pre-validated measures of alcohol use and dependence^19^. Social network size was elicited from a sequence of questions yielding the number of men who have sex with men, over the age of 18 living in Nairobi and met in person in the last two weeks.

Participants were offered HIV counselling and rapid testing following Kenyan HIV Testing Services (HTS) guidelines using two commercial rapid diagnostic kits (RDT: Determine Alere HIV 1/2 and First Response HIV 1–2.0)^20^. Blood specimens were tested for syphilis (TPHA/RPR), hepatitis B surface antigen and hepatitis C antibody (Mircrowell ELISA, Bios USA) and qualitative or quantitative HIV-1 PCR conditional on rapid test results (GeneXpert Qual or HIV-VL). Urine and rectal swabs were collected and tested for *Neisseria gonorrhoea* and *Chlamydia trachomatis* using PCR (GeneXpert CTNG).

HIV care continuum measures were based on CDC guidelines with a viral suppression threshold of <50 copies/ml^21^. Self-reported HIV status awareness and use of anti-retroviral therapy ART were collected both by computer-assisted survey and as part of HTS. Measures of linkage to care within 6 months of diagnosis and retention in care over the past 12 months were only elicited in the survey.

Persons living with HIV/AIDS (PLWHA) not reporting receipt of care were referred to government services for initiation of anti-retroviral therapy. HIV negative participants were referred for pre-exposure prophylaxis PrEP eligibility assessment. Treatment for other STIs was provided free and according to national guidelines. Condoms and water-based lubricants were freely available in the study clinic as was information about sexual risk reduction and other GBMSM/TP-affirming local sexual health services. Participants were compensated 500 Kenya shillings (∼USD $5) for completing study procedures, as approved by the ethics review board.

### Statistical methods

RDS diagnostics including visualisation of recruitment chains, convergence and seed dependence, and statistical assessment of recruitment homophily were analysed using the *rds* library for R version 3.4.0^22,23^. Crude and sample weighted estimates (RDS-II method and excluding seeds^23^) of the prevalence of sociodemographic and behavioural factors, lab-confirmed and self-reported STIs and HIV cascade measures (for PLWHA only) are presented in accordance with good practice^24^. Given evidence of under-reporting of status awareness and ART use in HTS and surveys alone (see supplementary materials), a composite cascade was derived combining both sources and treating any report of HIV awareness or treatment receipt as a positive response. Age and partner count quintiles among PLWHA were coded and used throughout for consistency.

Associations with HIV prevalence in the entire sample, and prevalence of detectable HIV viraemia among PLWHA only, were assessed using robust Poisson regression with a non-clustered sandwich estimator^25^ for an unbiased estimate of the prevalence ratio^26^. Multivariable models were specified including sociodemographic (model 1) or full (model 2) covariates associated with outcome at *p*<0.100. Given the bimodal distribution of viral load among PLWHA, comparisons between quantitative VL measures were limited to non-parametric significance testing (Kruskall-Wallis test) and distribution visualisation (Epanechnikov kernels). Analyses of association were not sample weighted, given the known risk of bias in applying network weights to multivariate analyses ^27^ and likely correlation of pertinent behavioural measures with social network degree. Less than 5% of covariate measures were missing and were included in models as dummy variables. Analyses were performed in Stata version 16.

This study was approved by the Kenya Medical Research Institute Scientific and Ethics Review Unit (KERMI/SERU/CGMR-C/CSC 044/3334), the University of Oxford, Oxford Tropical Research Ethics Committee (OxTREC 47-16) and London School of Hygiene & Tropical Medicine Human Research Ethics Committee (REF: 14144). All participants provided separate written informed consent to the questionnaire, sample collection and sample storage, and were able to withdraw from any portion of the study.

## Results

761 individuals presented to the study site with the intention of participation. 124 were ineligible due to fake or missing coupons, repeat attendance, intoxication or failure to meet inclusion criteria. Of the 637 individuals with confirmed eligibility, 29 declined participation during consent procedures. Of 608 recruits and 10 seeds completing informed consent, one participant declined blood testing and six declined rectal swabs. Four seeds accounted for 516 (84.9%) recruits. Depth of recruitment ranged from 1 to 19 waves per seed (median 7) (Appendix 1).

Table 1 shows the characteristics of enrolled participants. Median age was 24 years (IQR 21-29) with 38.2% between the ages of 18-22 years. Most participants reported having attended post-primary education, however a high proportion of participants reported being unemployed. A minority of participants reported a birthplace outside of Kenya, predominantly in neighbouring East African countries, in particular Uganda (n=90). Three-quarters of participants self-identified as gay or homosexual, and 15.0% self-identified as non-cisgender (predominantly transfeminine or female). Only 35.3% (30.9-39.9%, 229/580) reported having been in contact with community-based organisations targeting GBMSM/TP during the previous year.

**Table 1:**
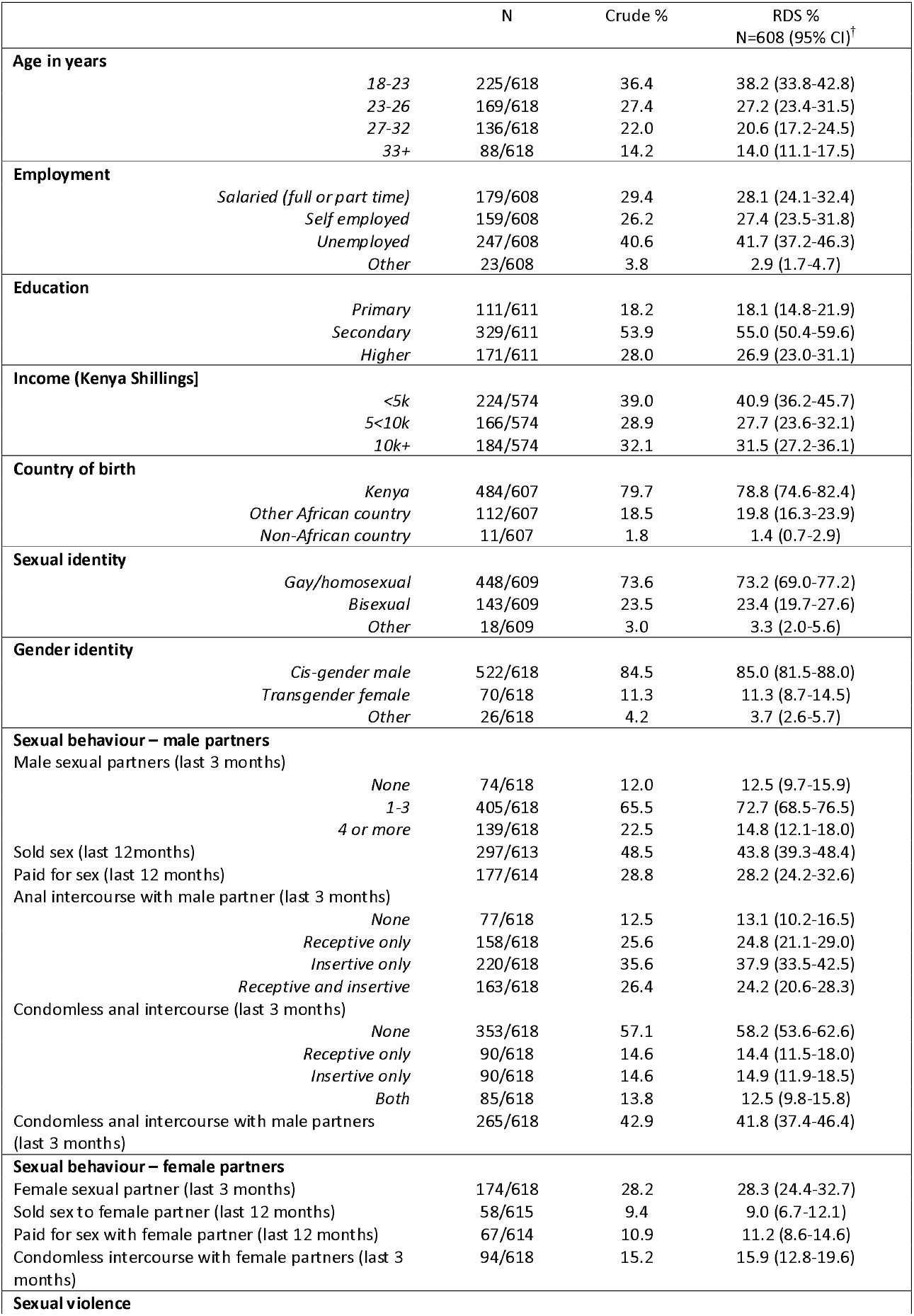
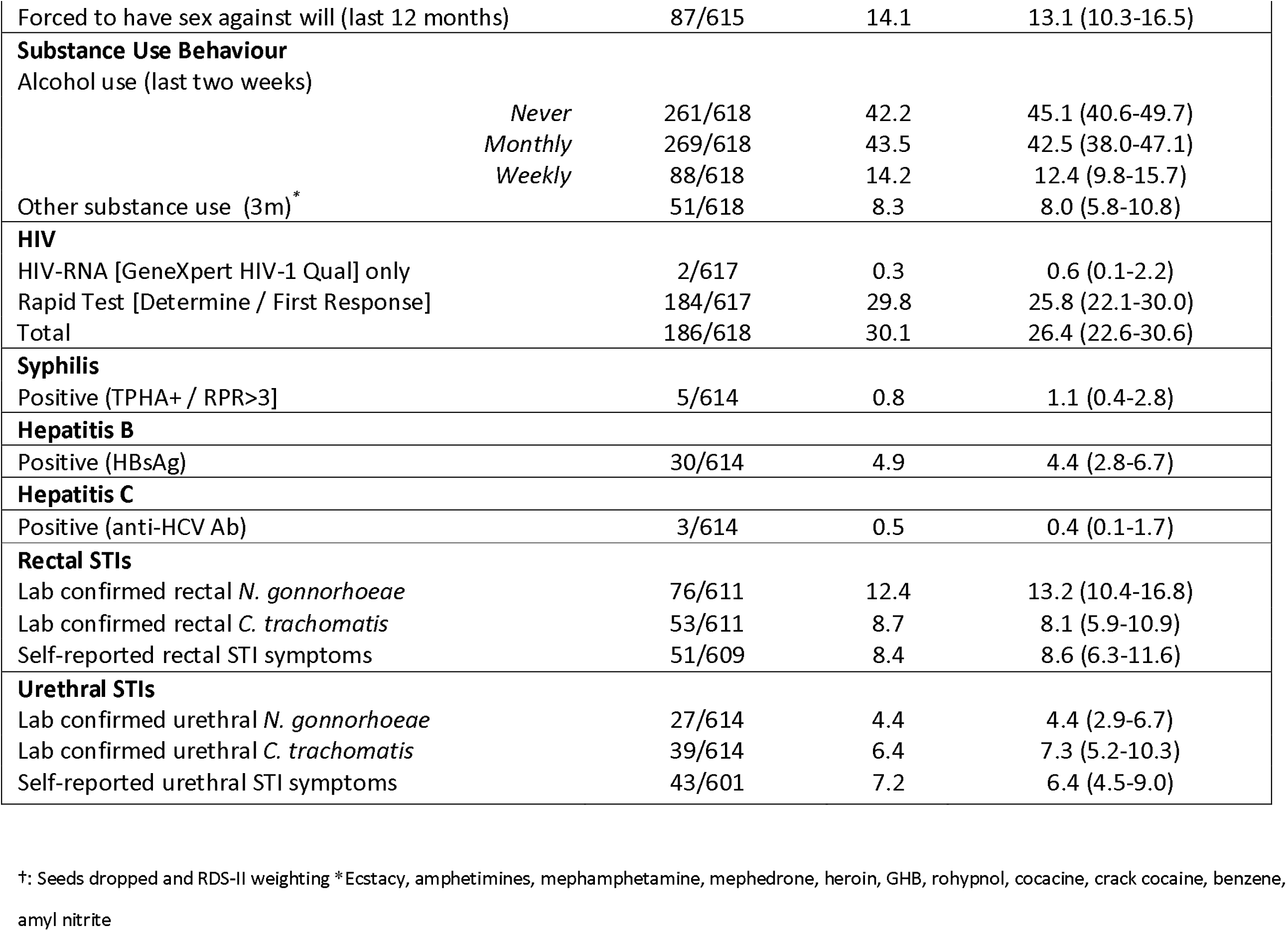
Sample sociodemographic characteristics. risk behaviour and sexually transmitted infections

|  | N | Crude % | RDS %<br>N=608 (95% CI)<sup>†</sup> |
| --- | --- | --- | --- |
| **Age in years** |  |  |  |
| 18-23 | 225/618 | 36.4 | 38.2 (33.8-42.8) |
| 23-26 | 169/618 | 27.4 | 27.2 (23.4-31.5) |
| 27-32 | 136/618 | 22.0 | 20.6 (17.2-24.5) |
| 33+ | 88/618 | 14.2 | 14.0 (11.1-17.5) |
| **Employment** |  |  |  |
| Salaried (full or part time) | 179/608 | 29.4 | 28.1 (24.1-32.4) |
| Self employed | 159/608 | 26.2 | 27.4 (23.5-31.8) |
| Unemployed | 247/608 | 40.6 | 41.7 (37.2-46.3) |
| Other | 23/608 | 3.8 | 2.9 (1.7-4.7) |
| **Education** |  |  |  |
| Primary | 111/611 | 18.2 | 18.1 (14.8-21.9) |
| Secondary | 329/611 | 53.9 | 55.0 (50.4-59.6) |
| Higher | 171/611 | 28.0 | 26.9 (23.0-31.1) |
| **Income (Kenya Shillings)** |  |  |  |
| <5k | 224/574 | 39.0 | 40.9 (36.2-45.7) |
| 5<10k | 166/574 | 28.9 | 27.7 (23.6-32.1) |
| 10k+ | 184/574 | 32.1 | 31.5 (27.2-36.1) |
| **Country of birth** |  |  |  |
| Kenya | 484/607 | 79.7 | 78.8 (74.6-82.4) |
| Other African country | 112/607 | 18.5 | 19.8 (16.3-23.9) |
| Non-African country | 11/607 | 1.8 | 1.4 (0.7-2.9) |
| **Sexual identity** |  |  |  |
| Gay/homosexual | 448/609 | 73.6 | 73.2 (69.0-77.2) |
| Bisexual | 143/609 | 23.5 | 23.4 (19.7-27.6) |
| Other | 18/609 | 3.0 | 3.3 (2.0-5.6) |
| **Gender identity** |  |  |  |
| Cis-gender male | 522/618 | 84.5 | 85.0 (81.5-88.0) |
| Transgender female | 70/618 | 11.3 | 11.3 (8.7-14.5) |
| Other | 26/618 | 4.2 | 3.7 (2.6-5.7) |
| **Sexual behaviour – male partners** |  |  |  |
| Male sexual partners (last 3 months) |  |  |  |
| None | 74/618 | 12.0 | 12.5 (9.7-15.9) |
| 1-3 | 405/618 | 65.5 | 72.7 (68.5-76.5) |
| 4 or more | 139/618 | 22.5 | 14.8 (12.1-18.0) |
| Sold sex (last 12 months) | 297/613 | 48.5 | 43.8 (39.3-48.4) |
| Paid for sex (last 12 months) | 177/614 | 28.8 | 28.2 (24.2-32.6) |
| Anal intercourse with male partner (last 3 months) |  |  |  |
| None | 77/618 | 12.5 | 13.1 (10.2-16.5) |
| Receptive only | 158/618 | 25.6 | 24.8 (21.1-29.0) |
| Insertive only | 220/618 | 35.6 | 37.9 (33.5-42.5) |
| Receptive and insertive | 163/618 | 26.4 | 24.2 (20.6-28.3) |
| Condomless anal intercourse (last 3 months) |  |  |  |
| None | 353/618 | 57.1 | 58.2 (53.6-62.6) |
| Receptive only | 90/618 | 14.6 | 14.4 (11.5-18.0) |
| Insertive only | 90/618 | 14.6 | 14.9 (11.9-18.5) |
| Both | 85/618 | 13.8 | 12.5 (9.8-15.8) |
| Condomless anal intercourse with male partners (last 3 months) | 265/618 | 42.9 | 41.8 (37.4-46.4) |
| **Sexual behaviour – female partners** |  |  |  |
| Female sexual partner (last 3 months) | 174/618 | 28.2 | 28.3 (24.4-32.7) |
| Sold sex to female partner (last 12 months) | 58/615 | 9.4 | 9.0 (6.7-12.1) |
| Paid for sex with female partner (last 12 months) | 67/614 | 10.9 | 11.2 (8.6-14.6) |
| Condomless intercourse with female partners (last 3 months) | 94/618 | 15.2 | 15.9 (12.8-19.6) |
| **Sexual violence** |  |  |  |

|  |  |  |  |  |
| --- | --- | --- | --- | --- |
| Forced to have sex against will (last 12 months) |  | 87/615 | 14.1 | 13.1 (10.3-16.5) |
| **Substance Use Behaviour** |  |  |  |  |
| Alcohol use (last two weeks) |  |  |  |  |
|  | *Never* | 261/618 | 42.2 | 45.1 (40.6-49.7) |
|  | *Monthly* | 269/618 | 43.5 | 42.5 (38.0-47.1) |
|  | *Weekly* | 88/618 | 14.2 | 12.4 (9.8-15.7) |
| Other substance use (3m)* |  | 51/618 | 8.3 | 8.0 (5.8-10.8) |
| **HIV** |  |  |  |  |
| HIV-RNA [GeneXpert HIV-1 Qual] only |  | 2/617 | 0.3 | 0.6 (0.1-2.2) |
| Rapid Test [Determine / First Response] |  | 184/617 | 29.8 | 25.8 (22.1-30.0) |
| Total |  | 186/618 | 30.1 | 26.4 (22.6-30.6) |
| **Syphilis** |  |  |  |  |
| Positive (TPHA+ / RPR>3] |  | 5/614 | 0.8 | 1.1 (0.4-2.8) |
| **Hepatitis B** |  |  |  |  |
| Positive (HBsAg) |  | 30/614 | 4.9 | 4.4 (2.8-6.7) |
| **Hepatitis C** |  |  |  |  |
| Positive (anti-HCV Ab) |  | 3/614 | 0.5 | 0.4 (0.1-1.7) |
| **Rectal STIs** |  |  |  |  |
| Lab confirmed rectal *N. gonorrhoeae* |  | 76/611 | 12.4 | 13.2 (10.4-16.8) |
| Lab confirmed rectal *C. trachomatis* |  | 53/611 | 8.7 | 8.1 (5.9-10.9) |
| Self-reported rectal STI symptoms |  | 51/609 | 8.4 | 8.6 (6.3-11.6) |
| **Urethral STIs** |  |  |  |  |
| Lab confirmed urethral *N. gonorrhoeae* |  | 27/614 | 4.4 | 4.4 (2.9-6.7) |
| Lab confirmed urethral *C. trachomatis* |  | 39/614 | 6.4 | 7.3 (5.2-10.3) |
| Self-reported urethral STI symptoms |  | 43/601 | 7.2 | 6.4 (4.5-9.0) |

†: Seeds dropped and RDS-II weighting \*Ecstasy, amphetamines, methamphetamine, mephedrone, heroin, GHB, rohypnol, cocaine, crack cocaine, benzene, amyl nitrite

On average, participants reported 2.99 (2.01-3.97) male sexual partners in the past three months. Male partner counts were higher among the 44% of participants who reported selling sex to men in the past year (mean 4.2 vs 2.0 different partners in the last 3 months, Kruskall-Wallis *p*<0.001). 49.0% (44.5-53.6) reported receptive anal intercourse in the past three months, and 28.3% (23.1-31.2) reported at least one episode of condomless receptive anal intercourse over that time. A significant proportion of participants reported sex against their will. Over a quarter of participants reported female sexual partners over that period and participants were similarly likely to have sold sex to, or purchased sex from, females. Among HIV negative participants, 59.2% (237/396 53.4-64.6%) reported HIV testing within the last 6 months and 4.4% (25/430 2.7-7.0%) reported current oral PrEP use.

186 participants tested HIV positive (crude 30.1%, RDS-II 26.4%). Two individuals were positive only on PCR testing, representing 2.1% (2/186, 0.5-8.2%) of PLWHA or 0.76% (2/426, 0.18-0.30%) of participants testing negative by the national RDT algorithm. Five participants had evidence of active syphilis infection, and hepatitis B and C prevalence was low. Laboratory confirmed rectal STIs were more prevalent than urethral STIs, and rectal GC was the most common site-specific STI. 82.2% confirmed rectal infections (90/106, 72.0-89.3%) and 82.3% confirmed urethral infections (49/60, 68.8-90.8) were asymptomatic on self-report.

Table 2 shows crude and adjusted variable associations with HIV status. Across models, increasing age was strongly associated with increasing HIV prevalence. In fully adjusted models HIV prevalence rose on average 6.4% per year of age (5.0-7.9%) *p*<0.001), from 13% among 18-22 year olds to 48.9% among those over 32 years of age. Participants reporting a birthplace outside Kenya but within Africa had less than half the HIV prevalence of Kenyan-born participants in all models. Transfeminine participants had a 50% higher prevalence than cisgender GBMSM after adjustment for sociodemographic factors, yet not after adjustment for behavioural factors. In crude analyses, HIV infection was associated with higher male partner counts, selling sex to men, and receptive anal intercourse. However, after adjustment, only reporting of recent receptive anal intercourse remained independently associated with HIV infection.

**Table 2:**
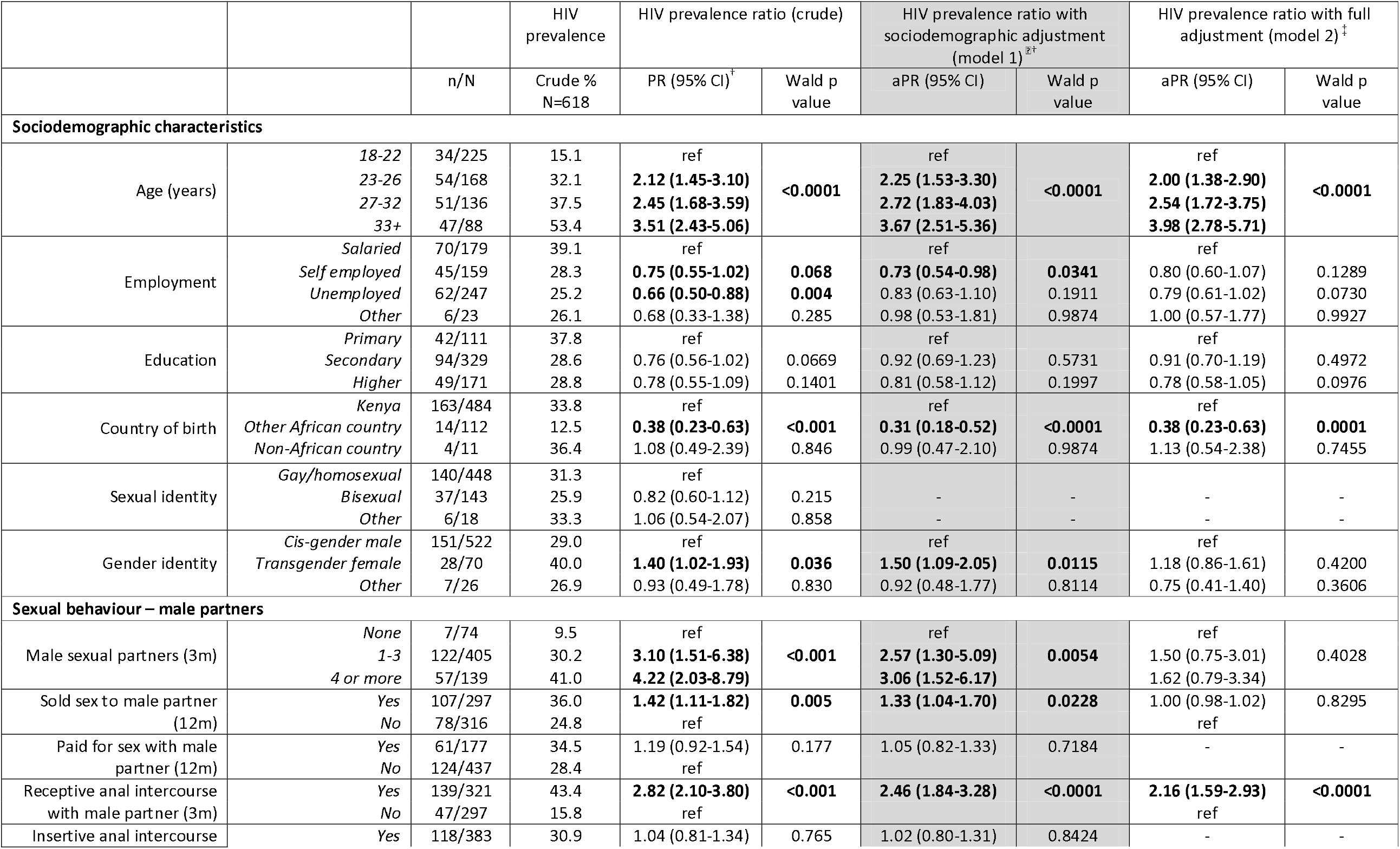
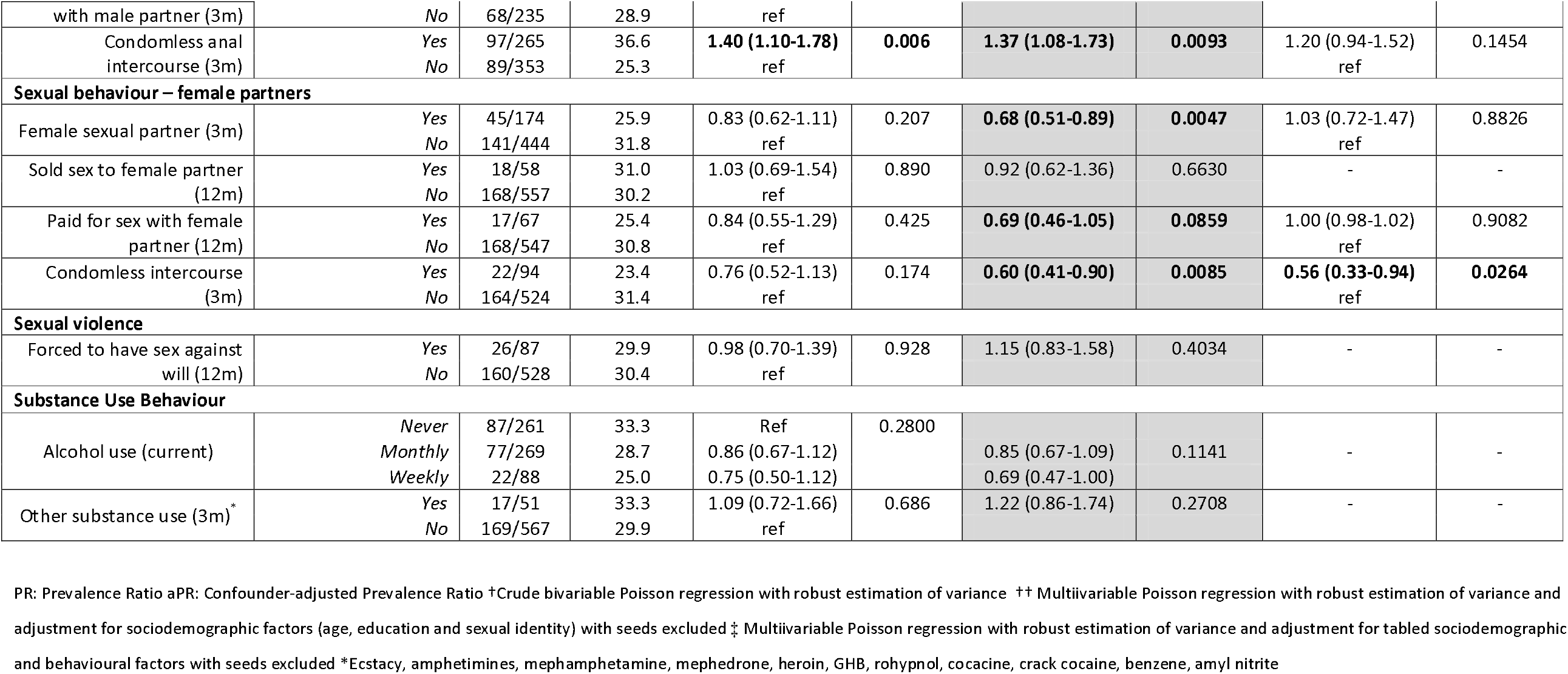
Associations with HIV status, GBMSM/TP, Nairobi 2017

Figure 1a shows the composite, RDS-II adjusted care cascade among participants with HIV infection (see Appendix 2 for cascades based on survey and HTS measures only). 97.9% (91.8-99.5%, RDS-II, n=184) were detected by the HTS regimen, 76.5% (68.2-83.3%, RDS-II, n=137)) reported status awareness and 65.3% (56.6-73.2%, RDS-II, n=129) reported currently receiving anti-retroviral therapy. 47.4% (38.9-56.0%), RDS-II, n=92) of PLWHA were virally supressed (<50 copies/ml). Median viral load was highest among two PCR positive participants with negative rapid tests (6.46 log_10_ copies/ml), and declined significantly by each progressive step across the care continuum (**Figure 1b**). Among 131 participants declaring receipt of HIV care, 61 (41.7 (31.9-52.2%) last received care in a community organisation, 44 (36.9% (27.4-47.6%) in a public hospital, and 26 (21.5% (14.1-31.3%) from a private provider.

**Figure 1:**
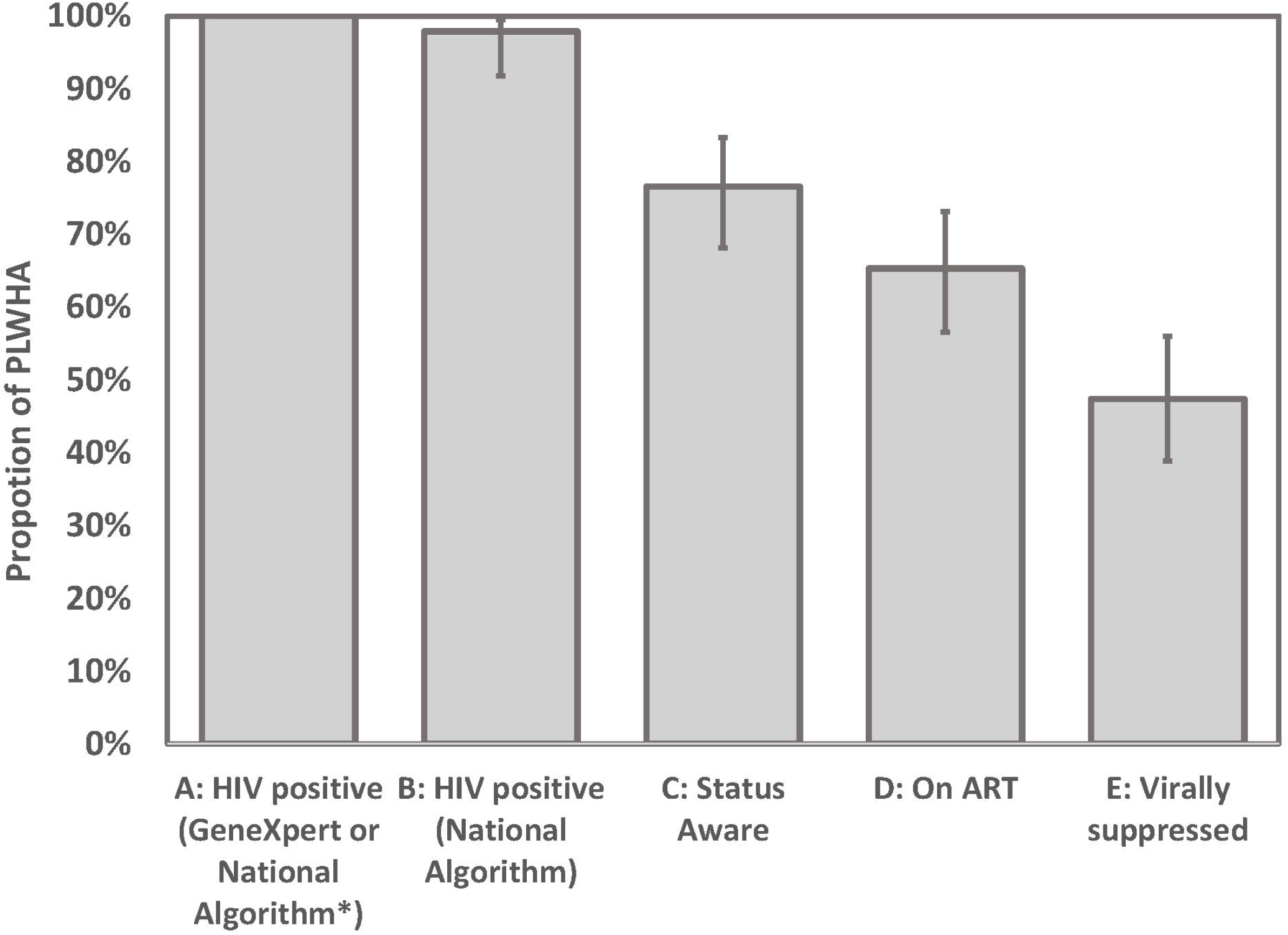
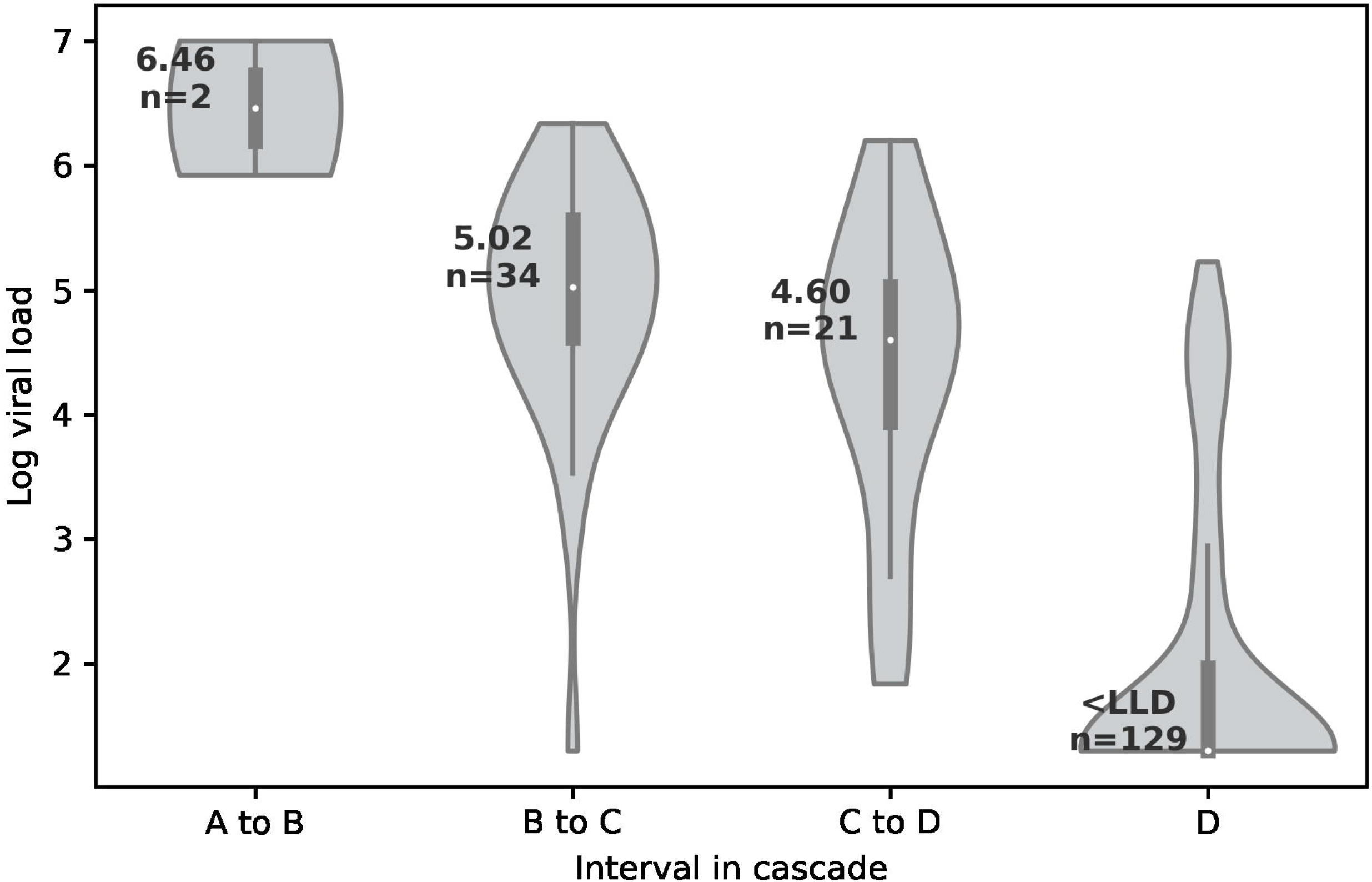
(a) Diagnosis and care cascade among GBMSM/TP living with HIV; (b) Log viral load median and distribution by level of diagnosis and care cascade engagement. Footnotes: **Footnote 1(a):** *Kenyan National HIV testing algorithm: Serial Determine Alere and First Response Rapid Diagnostic Tests Point estimates are RDS adjusted and exclude seeds. Error bars represent 95% confidence intervals **Footnote 2(b)** Intervals: A to B: HIV positive only on GeneXpert; B to C HIV positive on RDT but participant not status aware; C to D Participant reports status awareness but reports no current use of ART; D Participants reports current use of ART. Vertical bars represent interquartile range, white dots represent median log viral load. Median and category sample size stated in label. <LLD: below Lower Limit of Detection (40 copies/mm3). P-values from Kruskall-Wallis equality of populations rank test

Factors associated with detectable viraemia among PLHWA are reported in Table 3. A strong and significant inverse trend was apparent between increasing age and prevalence of detectable viraemia in both crude and adjusted models. On average, the prevalence of detectable HIV viraemia decreased by 4.2% per year of age (1.8-6.6%, test for linear trend *p*=0.0001). These trends were apparent across all metrics of the HIV care cascade (Figure 2a). Median log viral load among participants aged 18-22 was significantly higher than older age groups (4.44 vs 1.30 log_10_ copies/ml, Kruskall-Wallis *p*=0.0012, figure 2b), and both participants with acute HIV infections were within this youngest age-group. Increasing levels of education attendance were also associated with a declining level of viral detection among PLWHA, however this trend was not statistically significant. Correlates of prevalent HIV viraemia in the demographically adjusted model (model 1) were payment for sex in the last three months (with either male or female partners) and recent condomless anal intercourse with female partners, whilst there was an inverse association with recently selling sex to male partners.

**Table 3:** Associations with detectable VL among participants living with HIV, GBMSM/TP, Nairobi 2017

|  |  |  | Prevalence of detectable viral load >50 copies/ml | Viral detection prevalence ratio (crude) <sup>†</sup> |  | Viral detection prevalence ratio with sociodemographic adjustment (model 1) <sup>‡†</sup> |  | Viral detection ratio with full adjustment (model 2) <sup>‡</sup> |  |
| --- | --- | --- | --- | --- | --- | --- | --- | --- | --- |
|  |  | n/N | Crude % N=186 | PR (95% CI) | Wald p value | aPR (95% CI) | Wald p value | aPR (95% CI) | Wald p value |
| **Sociodemographic characteristics** |  |  |  |  |  |  |  |  |  |
| Age (years) | 18-22 | 25/34 | 73.5 | ref |  | ref |  | ref |  |
|  | 23-26 | 29/54 | 53.7 | **0.73 (0.53-1.01)** | **0.0020** | **0.74 (0.53-1.04)** | **0.0052** | **0.84 (0.61-1.16)** | **0.0103** |
|  | 27-32 | 24/51 | 47.1 | **0.64 (0.45-0.92)** |  | **0.65 (0.45-0.94)** |  | **0.71 (0.50-1.02)** |  |
|  | 33+ | 16/47 | 34.0 | **0.44 (0.28-0.70)** |  | **0.46 (0.29-0.74)** |  | **0.46 (0.29-0.74)** |  |
| Employment | Salaried | 37/70 | 72.9 | Ref |  | - | - | - | - |
|  | Self employed | 21/45 | 46.7 | 0.89 (0.61-1.32) | 0.569 | - | - | - | - |
|  | Unemployed | 33/62 | 53.2 | 1.02 (0.73-1.41) | 0.911 | - | - | - | - |
|  | Other | 2/6 | 33.3 | 0.64 (0.20-2.03) | 0.447 | - | - | - | - |
| Education | Primary | 27/42 | 64.3 | ref |  | ref | ref | ref |  |
|  | Secondary | 47/94 | 50.0 | 0.79 (0.58-1.08) | 0.1334 | 0.77 (0.56-1.05) | 0.1015 | 0.81 (0.60-1.08) | 0.1597 |
|  | Higher | 20/49 | 40.8 | **0.64 (0.43-0.97)** | **0.0355** | **0.64 (0.42-0.97)** | **0.0371** | 0.68 (0.46-1.00) | 0.0501 |
| Country of birth | Kenya | 86/163 | 52.8 | ref |  | - | - | - | - |
|  | Other African country | 6/14 | 42.9 | 0.82 (0.44-1.52) | 0.524 | - | - | - | - |
|  | Non-African country | 1/4 | 25.0 | 0.48 (0.09-2.63) | 0.395 | - | - | - | - |
| Sexual identity | Gay/homosexual | 77/140 | 55.0 | ref |  | ref |  | ref |  |
|  | Bisexual | 13/37 | 35.1 | 0.66 (0.42-1.05) | 0.083 | 0.78 (0.50-1.22) | 0.2803 | 0.68 (0.45-1.03) | 0.0646 |
|  | Other | 4/6 | 66.7 | 1.23 (0.68-2.21) | 0.496 | 1.31 (0.78-2.18) | 0.3065 | 1.33 (0.80-2.21) | 0.2741 |
| Gender identity | Cis-gender male | 76/151 | 50.3 | ref |  | - | - | - | - |
|  | Transgender female | 16/28 | 57.1 | 1.14 (0.80-1.64) | 0.467 | - | - | - | - |
|  | Other | 2/7 | 28.6 | 0.57 (0.17-1.87) | 0.355 | - | - | - | - |
| **Sexual behaviour – male partners** |  |  |  |  |  |  |  |  |  |
| Male sexual partners (3m) | None | 3/7 | 42.9 | ref |  | ref |  | - |  |
|  | 1-3 | 71/122 | 58.2 | **1.35 (0.56-3.23)** | **0.0336** | 1.04 (0.48-2.24) | 0.1202 | - | - |
|  | 4 or more | 20/57 | 35.1 | **0.81 (0.32-2.05)** |  | 0.68 (0.29-1.58) |  | - |  |
| Sold sex to male partner (12m) | Yes | 46/107 | 43.0 | **0.69 (0.52-0.91)** | **0.010** | **0.66 (0.50-0.86)** | **0.0028** | **0.55 (0.41-0.75)** | **0.0001** |
|  | No | 48/78 | 61.5 | ref |  | ref |  | ref |  |
| Paid for sex with male partner (12m) | Yes<br>No | 37/61<br>57/124 | 60.7<br>46.0 | **1.28 (0.96-1.70)**<br>Ref | **0.090** | 1.44 (1.10-1.88)<br>ref | 0.0084 | **1.72 (1.25-2.35)**<br>ref | **0.0008** |
| Receptive anal intercourse with male partner (3m) | Yes<br>No | 73/139<br>21/47 | 52.5<br>44.7 | 1.25 (0.86-1.83)<br>ref | 0.242 | 1.04 (0.72-1.50)<br>ref | 0.8420 | -<br>- | -<br>- |
| Insertive anal intercourse with male partner (3m) | Yes<br>No | 59/118<br>35/68 | 50.0<br>51.5 | 0.96 (0.72-1.29)<br>ref | 0.803 | 1.07 (0.81-1.43)<br>ref | 0.6192 | -<br>- | -<br>- |
| Condomless anal intercourse (3m) | Yes<br>No | 56/97<br>38/89 | 57.7<br>42.7 | **1.35 (1.00-1.81)**<br>ref | **0.051** | 1.30 (0.97-1.74)<br>ref | 0.0740 | 1.24 (0.95-1.63)<br>ref | 0.1166 |
| **Sexual behaviour – female partners** |  |  |  |  |  |  |  |  |  |
| Female sexual partner (3m) | Yes<br>No | 24/45<br>70/141 | 53.3<br>49.7 | 1.08 (0.78-1.49)<br>ref | 0.630 | 1.26 (0.94-1.67)<br>ref | 0.1164 | -<br>- | -<br>- |
| Sold sex to female partner (12m) | Yes<br>No | 9/18<br>85/168 | 50.0<br>50.6 | 0.99 (0.61-1.62)<br>ref | 0.981 | 0.96 (0.65-1.41)<br>ref | 0.8344 | -<br>- | -<br>- |
| Paid for sex with female partner (12m) | Yes<br>No | 14/17<br>80/168 | 82.4<br>47.6 | **1.74 (1.33-2.29)**<br>ref | **<0.001** | **1.64 (1.26-2.11)**<br>ref | **0.0002** | 1.22 (0.90-1.66)<br>ref | 0.1912 |
| Condomless intercourse 3m | Yes<br>No | 14/22<br>80/164 | 63.6<br>48.8 | 1.31 (0.92-1.87)<br>ref | 0.132 | **1.62 (1.19-2.21)**<br>ref | **0.0023** | 1.37 (0.96-1.95)<br>ref | 0.0789 |
| **Sexual violence** |  |  |  |  |  |  |  |  |  |
| Forced to have sex against will (12m) | Yes<br>No | 11/26<br>83/160 | 42.3<br>51.9 | 0.82 (0.51-1.32)<br>ref | 0.413 | 0.83 (0.55-1.28)<br>ref | 0.4022 | -<br>- | -<br>- |
| **Substance Use Behaviour** |  |  |  |  |  |  |  |  |  |
| Alcohol use (current) | Never<br>Monthly<br>Weekly | 47/87<br>3/77<br>10/22 | 54.0<br>48.1<br>45.5 | ref<br>0.90 (0.66-1.22)<br>0.85 (0.52-1.40) | 0.7032 | ref<br>0.94 (0.70-1.26)<br>0.90 (0.56-1.45) | 0.8611 | -<br>-<br>- | -<br>-<br>- |
| Other substance use (3m)* | Yes<br>No | 11/17<br>83/169 | 64.7<br>49.1 | 1.27 (0.84-1.92)<br>ref | 0.250 | 1.28 (0.85-1.94)<br>ref | 0.2425 | -<br>- | -<br>- |

PR: Prevalence Ratio aPR: Confounder-adjusted Prevalence Ratio †Crude bivariable Poisson regression with robust estimation of variance \*\* Multivariable Poisson regression with robust estimation of variance and adjustment for sociodemographic factors (age, education and sexual identity) with seeds excluded ‡ Multivariable Poisson regression with robust estimation of variance and adjustment for tabled sociodemographic and behavioural factors with seeds excluded \*Ecstasy, amphetamines, methamphetamine, mephedrone, heroin, GHB, rohypnol, cocaine, crack cocaine, benzene, amyl nitrite

**Figure 2:**
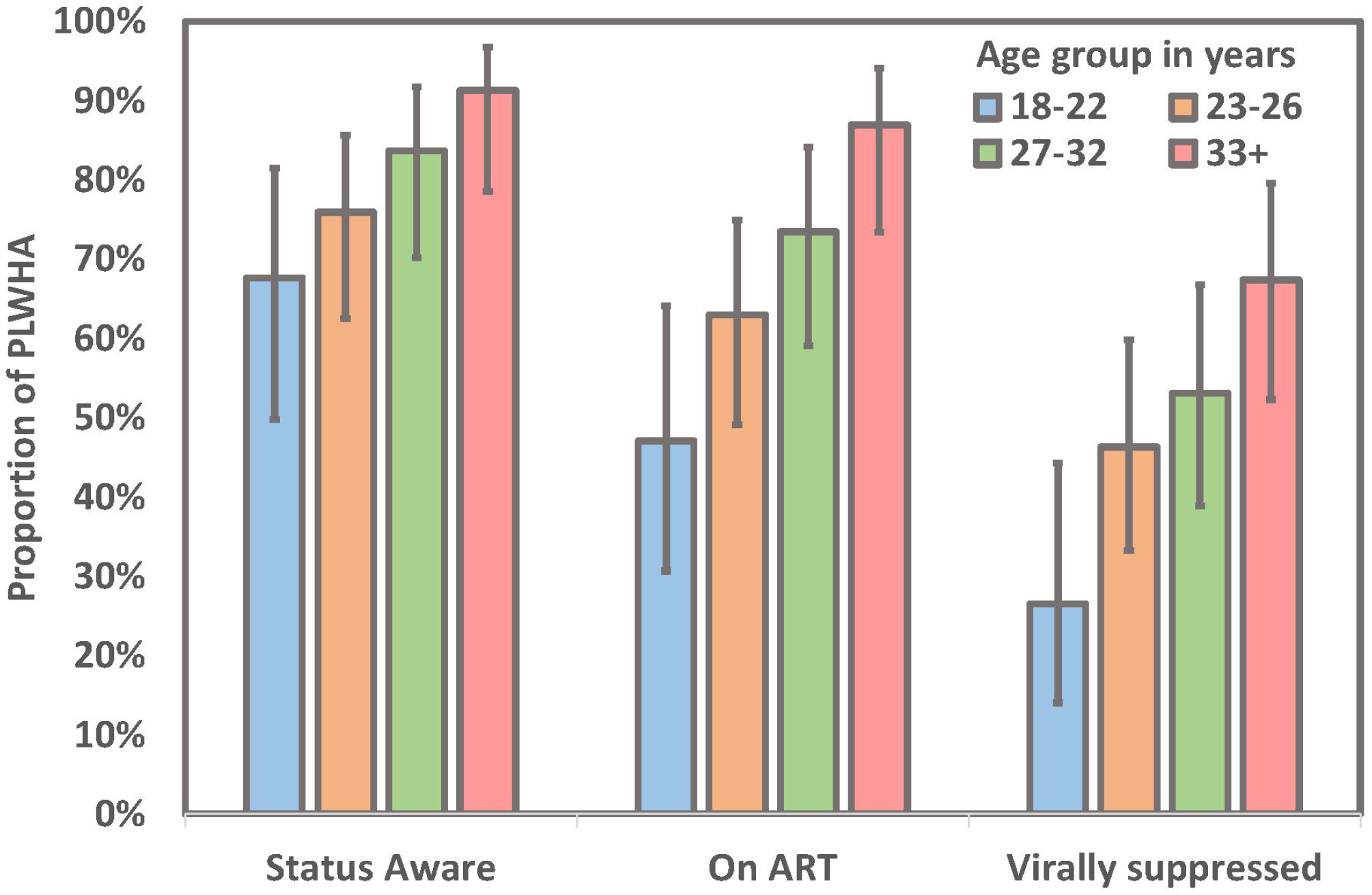
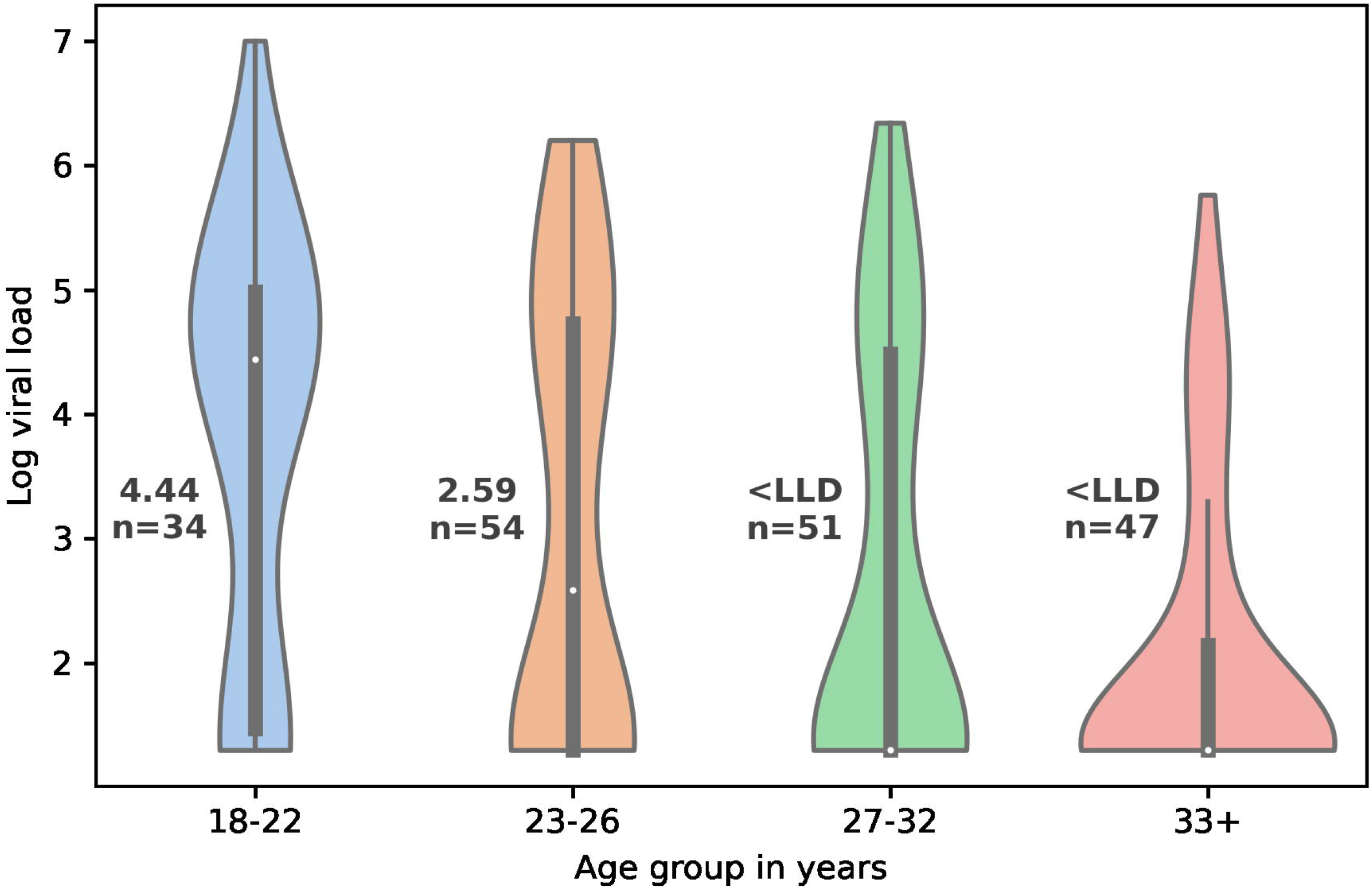
(a) HIV care cascade measures by age group; (b) Log viral load median and distribution by age group. **Footnote 2(a)** Point estimates are unadjusted for sampling strategy and exclude seeds Error bars represent 95% confidence intervals **Footnote 2(b)** Vertical bars represent interquartile range, white dots represent median viral load (also stated in label) <LLD: below Lower Limit of Detection (40 copies/mm3)

## Discussion

Over a quarter of GBMSM and TP in Nairobi now live with HIV infection. These estimates are higher than previous estimates from the same city (18.2%^13^) and compared to convenience samples elsewhere in Kenya (19.8% Malindi 2010^28^; 16.6% Kisumu 2015^29^). Extrapolation of the observed proportion with evidence of acute/early HIV infection not detectable by 4^th^ generation testing (estimated 14 day window period) suggests an annual HIV incident risk of 15% (4-58%). Persistently high HIV/STI risk among is consistent with high reported behavioural and biological acquisition risks that have not improved over time^13^: over 40% of GBMSM/TP report recent condomless anal intercourse and transactional partnerships, and a high proportion have co-occurring and asymptomatic STIs. Anti-retroviral prevention uptake remains poor and whilst the national PrEP programme was in the process of deployment during this study, subsequent evaluation since confirms inadequate uptake and persistence among GBMSM^30^.

However, this study does highlight significant progress in reaching key populations with HIV testing and care. We estimate that three-quarters of GBMSM/TP living with HIV in Nairobi are aware of their status and nearly half have been supported to achieve viral suppression, analogous to 77-85-73 against UNAIDS targets. This cascade compares favourably to GMSMS/TP cascades data elsewhere in sub-Saharan Africa (18-53-76^9^) as well as to that reported in global self-reported surveys (NA-82-58)^31^. This is by no means a small achievement of HIV programming within a societal context of homophobic discrimination and criminalisation of same sex behaviour^6^ and represents marked improvements in access to HIV care that will directly translate into better health outcomes for GBMSM and TP living with HIV. However, cascades fall behind those for PLWH in the Kenyan general population (80-96-91 in 2017)^12^ and for GBMSM and transgender in high income settings^32^.

Inequalities in coverage of HIV diagnosis and care were principally driven by age. We observed strong positive associations between increasing age and virological suppression, as well as other metrics of the care cascade. Median viral load was 3.14 log higher among participants age 18-22 living with HIV than older GMSM/TP (4.44 v 1.30 respectively, *p=*0.0022), reflecting both lower status awareness and care engagement in addition to higher HIV incident risk in the youngest age group. The observation that HIV prevalence was 13% among 18-22 year old GBMSM/TP suggests that risk begins earlier in adolescence when prevention and care may be even less accessible. Although comparable evidence is scarce from elsewhere in sub Saharan Africa, Ramadhani reports higher HIV risk behaviour and incidence, yet lower healthcare engagement, status awareness and virological suppression among 16-19 year old Nigerian GBMSM/TP^33^

The WHO highlight the need for national responses to be acceptable to young key populations^34^, and our findings suggest a focus on GBMSM/TP youth is not only overdue but will be essential to the overall success of Kenyan key population HIV response. Improving accessibility to youth may require redress of structural barriers to service access, such as age-based consent criteria, training of staff to recognise additional needs of young MSM, but must also account for the prospect that young MSM will be sceptical of confidentiality and safety of healthcare settings^35^. Pettifor proposes that services for adolescent and young MSM need to be targeted and holistic, given the complex and concurrent challenges of conceptualising HIV risk and prevention during a period of personal biological and psychological change, and often alongside stressors related to acceptance and disclosure of sexual or gender identity to family and friends^35^.

Our findings also suggest that improved diagnostics could complement both HIV prevention and care for GBMSM/TP in Nairobi. A small but significant proportion of GBMSM/TP were identified with prevalent acute/early HIV infection accompanied by high viral loads, and undetected by current national testing practices. In addition we found a high proportion of GBMSM/TP with asymptomatic, urethral and rectal STIs, well recognised as a co-factor in HIV transmission^36^. Laboratory capacity for STI diagnosis remains limited and expensive in Kenya, therefore most providers, especially community-based organisations, rely solely upon syndromic management. Our findings concur with others in suggesting such approaches alone have unacceptably poor diagnostic performance^37,38^. The decreasing complexity and cost of point-of-care PCR technologies should encourage policy makers to re-evaluate the cost effectiveness of providing access to PCR-based HIV and STI diagnostics particularly in community settings^39^.

A key strength of the study was the population representative design which avoided many of the biases intrinsic to studies conducted solely among GBMSM/TP already engaged with service providers. RDS diagnostics suggest convergence on all main demographic measures, and these compared closely to a previous study of the same design in Nairobi^13^. Limitations of the study include the cross-sectional design (precluding examination of causal direction of correlates) and the reliance on self-reported measures of behaviours and service uptake that are potentially subject to memory error and social desirability bias. Foremost among these was differential under-reporting of status awareness and anti-retroviral use in surveys and with care providers. This phenomenon has been reported by a few other population-based studies and has the potential to significantly distort interpretation of cascade measures^40,41^ and underscores the need for verification of self-reported measures wherever possible.

In summary, coverage of HIV care for GBMSM and transgender persons living with HIV in Nairobi is close to that achieved in the general population and reflects the inclusive approach of the national HIV/AIDS strategy in Kenya. However, ending AIDS for key populations demands even better access to care, a re-energised PrEP response, and relevant HIV and STI diagnostics available wherever GBMSM/TP feel safe seeking these services. Going forward policy makers must now seek to understand and address the sexual health service preferences of adolescent and younger key populations in order to address existing inequalities in access to diagnosis and care services.

## Supporting information

Appendix 1: Respondent Driven Sampling recruitment diagnostics

Appendix 2: Sensitivity analysis of source of cascade definitions and viral load cutoffs

## Data Availability

Data from this study has not been deposited publicly because of the potential risk of deductive disclosure that may arise from individual data needed for valid analysis of the data, and the potential individual and social harms that may arise from such disclosure in a context of criminalisation and stigmatisation. However all authors aim to make the data underlying the findings of the study available for legitimate research purposes, and requests will be considered by the London School of Hygiene and Tropical Medicine Research Operations Office Data Management lead. The request must specify the purpose of research, the list of required variables, and if personally identifiers or sensitive data are sought, specify measures to maintain information security and governance that will be applied in storage, handling and reporting the data.

## Data Availability

Code is publicly available on GitHub (https://github.com/bnwolford/FHiGR_score) and summary statistics are made available in the manuscript. Individual level data is not available for participant privacy.

## Acknowledgements

We would like to acknowledge and thank the commitment of study participants, and are grateful to our community partner organisations: the Gay and Lesbian Coalition of Kenya (GALCK); Ishtar MSM and Health Options for Young Men with AIDS (HOYMAS) for their support of study procedures and in dissemination of findings. We thank our administrative, counselling, clinical and laboratory staff at the TRANSFORM clinic and Partners for Health and Development for Africa (PHDA) for their hard work and dedication.

This study was funded by Evidence for HIV Prevention in Southern Africa (EHPSA), a UK aid programme managed by Mott MacDonald (award reference MM/EHPSA/WHC/0116029). The funder of the study had no role in study design, data collection, data analysis, data interpretation of writing of the report.

## Author contributions

ADS contributed to designing the study and data collection instruments, carried out quantitative analyses and wrote the first draft of the manuscript; AB contributed to conceiving and designing the study and data collection instruments and drafting of the manuscript; JK and RK contributed to designing the study and data collection instruments, implementation of study procedures, and commented on the manuscript. EI, MK, PM, HB and CN contributed to the implementation and operation of study procedures. PW and EF contributed to conceiving and designing the study and data collection instruments and commented on the manuscript. All authors approved the final draft.

