## Appendix 1: Respondent Driven Sampling recruitment diagnostics for "Age-dependent inequalities in HIV/STI burden and care receipt among men and transgender persons who have sex with men in Nairobi"

In line with guidance[1] we conducted monitoring of the RDS recruitment every two weeks and made recommendations to the data collection team on the basis of findings. We examined RDS recruitment trees, return of coupons, assessed sample composition by sociodemographic characteristics, and examined convergence on key outcomes overall and by seed (assessed for potential 'bottlenecks' in which recruitment gets 'stuck' within sub-groups).

#### Findings

##### Waves and seeds

Ten seed participants recruited a total of 608 participants, though 512/608, 84% of all participants, came from 4 seeds of between 14 and 19 waves, Table A1. The largest recruitment chain of Seed 1 recruited 143/608 (23.5%) of participants, whilst Seed 3 had the largest number of recruitment waves, Table A2.

**Table A1: Number of participants recruited by wave**

| Wave | 0 | 1 | 2 | 3 | 4 | 5 | 6 | 7 | 8 | 9 | 10 | 11 | 12 |
|---|---|---|---|---|---|---|---|---|---|---|---|---|---|
| No. Participants | 10 | 19 | 29 | 43 | 57 | 69 | 80 | 64 | 66 | 49 | 37 | 29 | 24 |

| Wave | 13 | 14 | 15 | 16 | 17 | 18 | 19 |
|---|---|---|---|---|---|---|---|
| No. Participants | 12 | 12 | 4 | 6 | 6 | 1 | 1 |

**Table A2: Number of participants and recruitment waves recruited via each seed**

| Seed | Number of participants (%) | Waves |
|---|---|---|
| 1 | 143 (23.5%) | 17 |
| 2 | 17 (2.8%) | 6 |
| 3 | 139 (22.9%) | 19 |
| 4 | 139 (22.9%) | 15 |
| 5 | 39 (6.4%) | 8 |
| 6 | 4 (0.7%) | 2 |
| 7 | 91 (15.0%) | 14 |
| 8 | 22 (3.6%) | 6 |
| 9 | 2 (0.3%) | 1 |
| 10 | 12 (2.1%) | 4 |

##### Coupon receipt

96.9% of participants received a coupon from a close friend or friend. Only 2 (0.4%) of participants reported that they received their coupon from a stranger, which violates the RDS assumption that participants receive a coupon from someone within their social network (and the recruitment instructions). Consistent with whom they received a coupon from, the majority of participants reported that they received it from or near their home. There were 17 participants (2.8%) who reported receiving their coupon outside the study clinic which could imply distributions to individuals who just happened to be passing rather than distribution to an individual's social network, but this is not certain.

A minority of 45/600 (7.5%) of participants reported that they had received more than one offer of a coupon, which suggests that recruitment did not get stuck within a small group trying to recruit each other and did not saturate the target population.

**Table A3: Characteristics of coupon receipt reported by participants**

| | | n | % |
|---|---|---|---|
| **Relationship to person from whom received coupon (n=576)** | | | |
| Close friend | | 292 | 50.7 |
| Friend | | 266 | 46.2 |
| Acquaintance | | 14 | 2.4 |
| Stranger | | 2 | 0.4 |
| Other | | 2 | 0.4 |
| **Where received coupon (n=598)** | | | |
| At/near home | | 301 | 50.3 |
| At/near work | | 76 | 12.7 |
| On street | | 86 | 14.4 |
| Bar/club | | 96 | 16.1 |
| Outside the study clinic | | 17 | 2.8 |
| Other | | 22 | 3.7 |
| **Apart from the person who gave you the coupon you brought today has anyone else tried to give you a coupon? (n=600)** | | | |
| No | | 555 | 92.5 |
| Yes | | 45 | 7.5 |
| | **How many times?** | | |
| | 1 | 19 | 48.7 |
| | 2 | 15 | 38.5 |
| | 4 | 1 | 2.6 |
| | 5 | 2 | 5.1 |
| | 7 | 1 | 2.6 |
| | 11 | 1 | 2.6 |

#### Convergence of Estimates

Each convergence plot shows the cumulative RDS-II weighted proportion of the population estimate as the sample recruitment progressed. We examined key sociodemographic characteristics, HIV prevalence and viral suppression.

#### Figure A2

##### a. Age: proportion 25 years or more

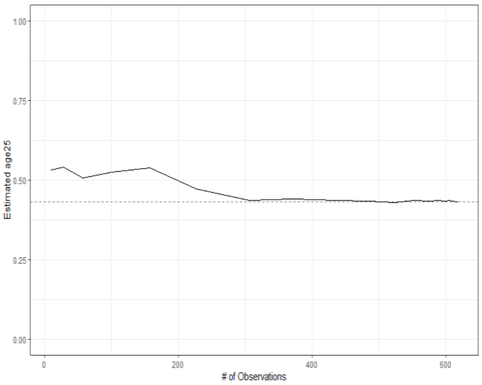


##### e. Gender identity

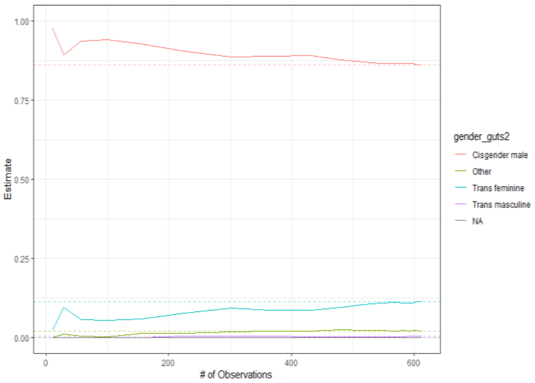


##### b. Educational attendance

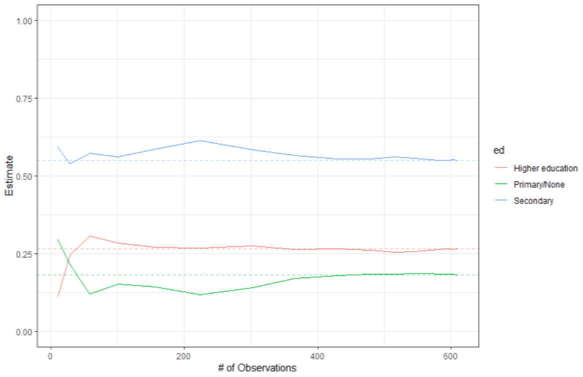


##### f. HIV: proportion positive

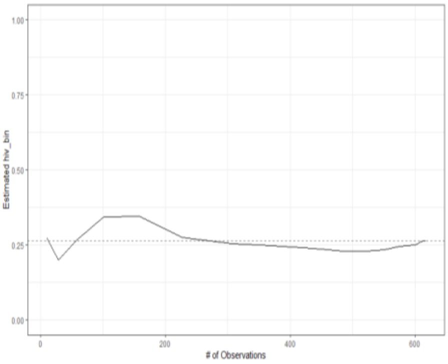


##### c. Employment

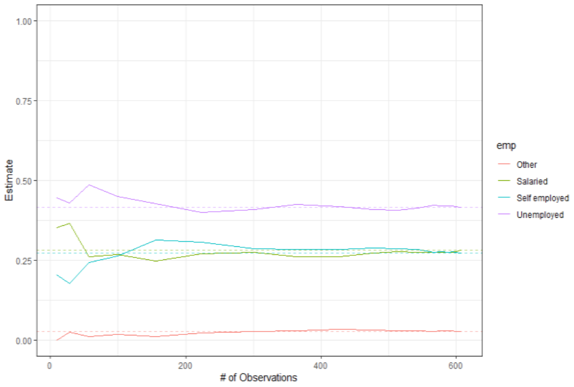


##### g. Viral suppression: proportion of PLWHA

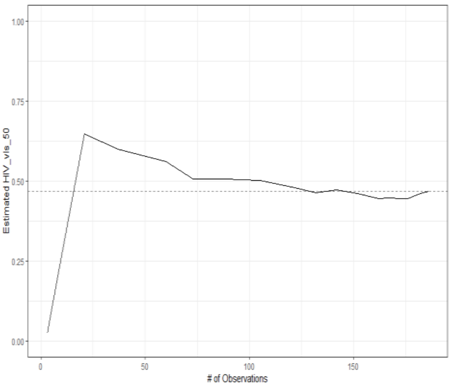


##### d. Sexual identity

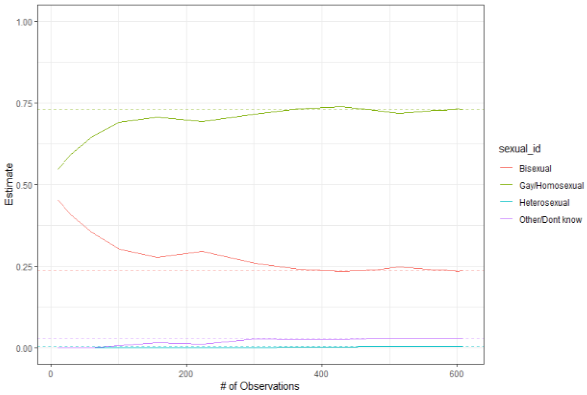

#### Bottlenecks

We show the bottleneck plot for basic demographic features only to limit the potential for deductive disclosure of seed identity.

##### Figure A3a. Age: proportion 25 years or more

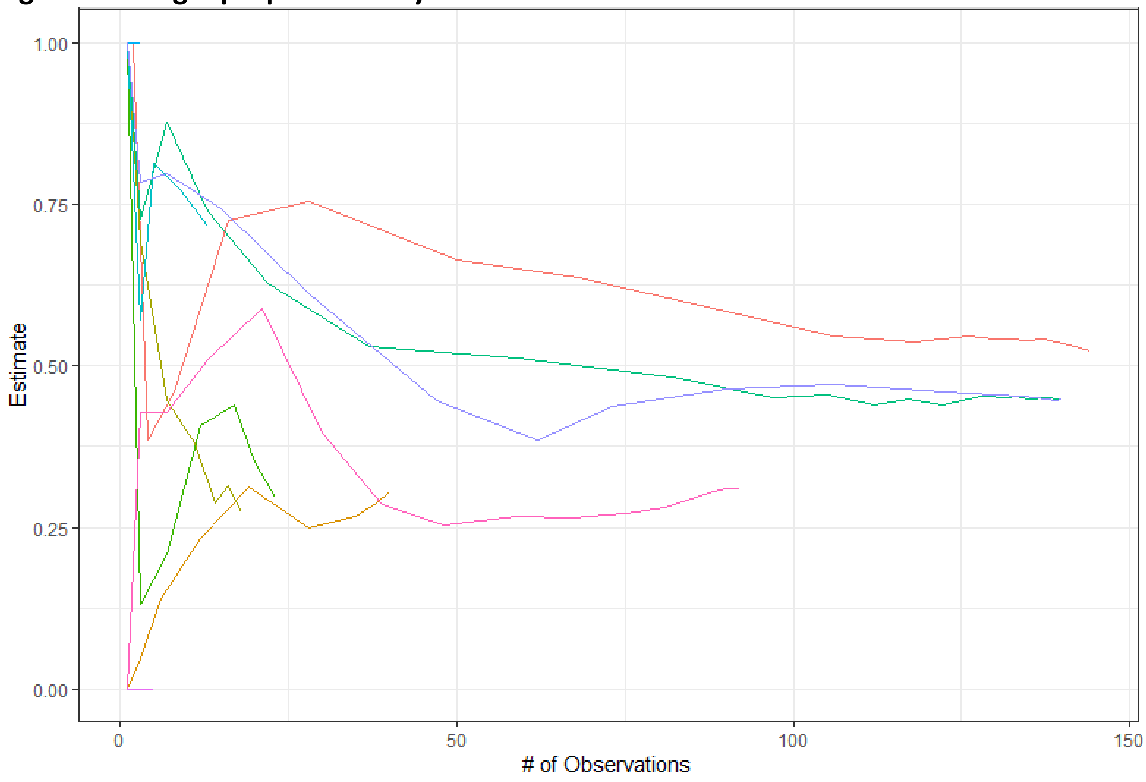


##### Figure A3b. Educational attendance: proportion attended higher education

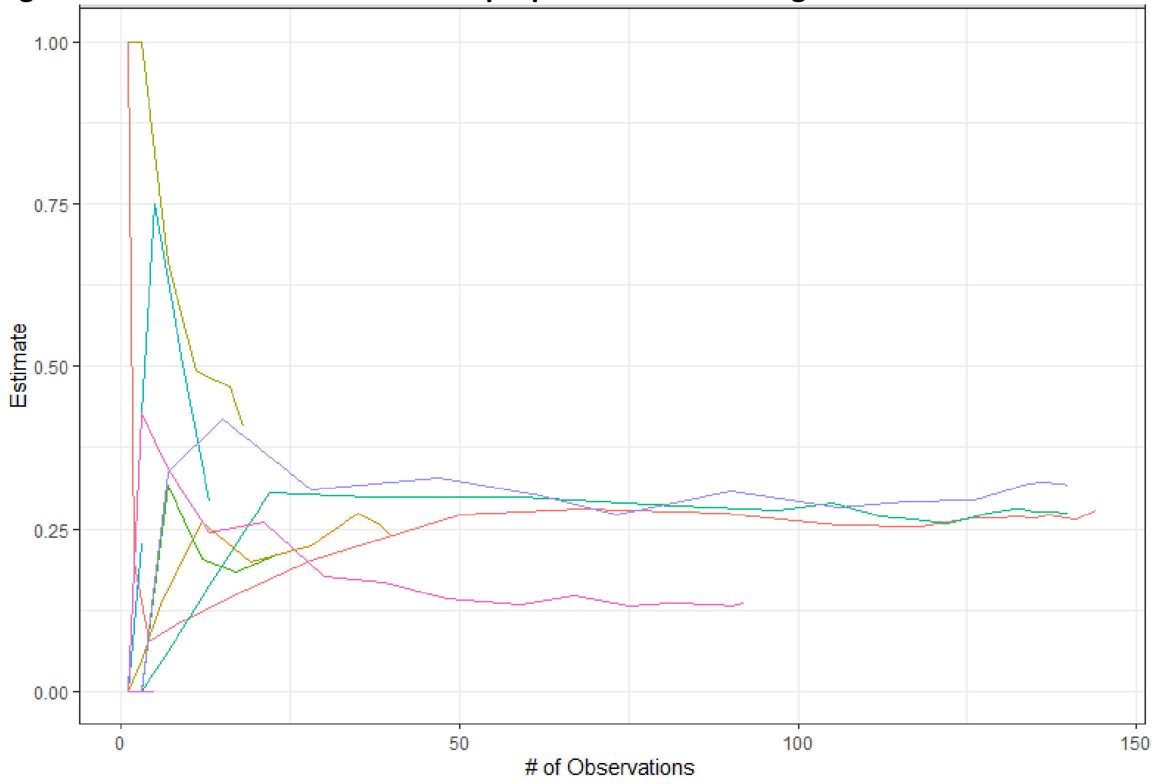

**Figure A3c. Current employment: proportion unemployed**

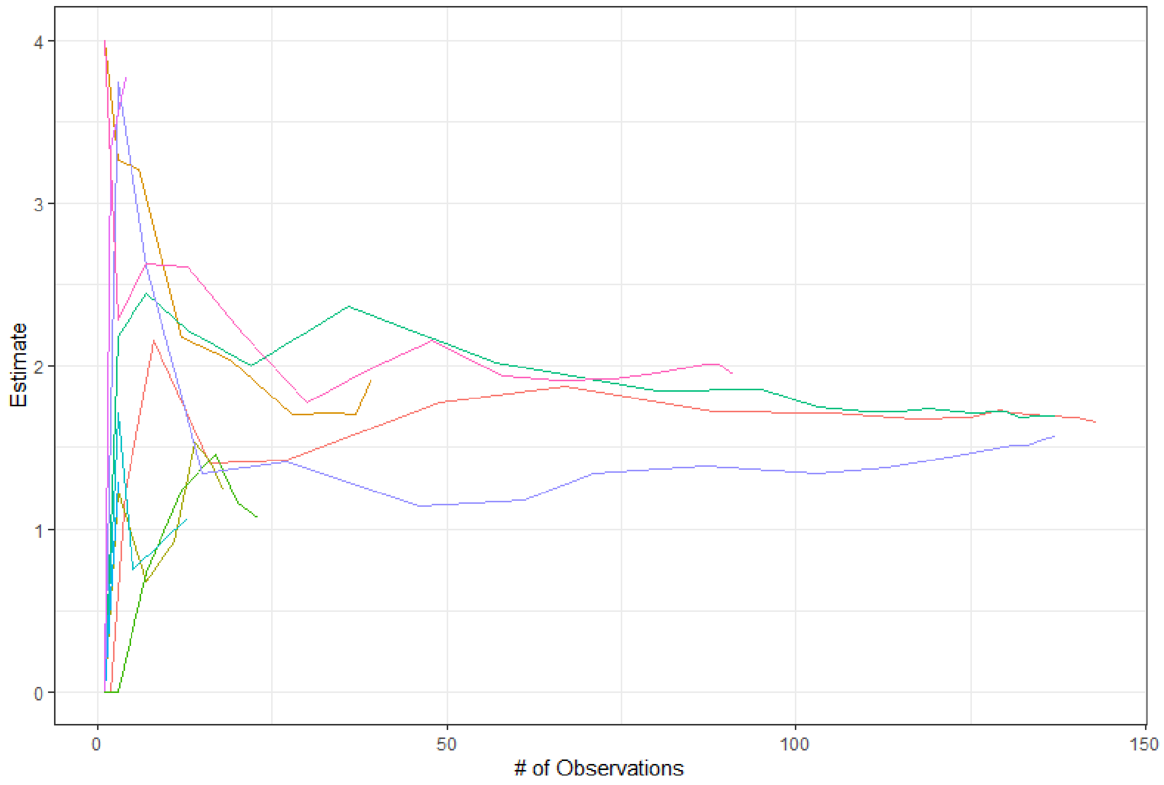


Whilst the sample achieved equilibrium on age (Figure A2a) and the trajectory of individual recruitment chains were heading for convergence, there was some evidence for persisting seed dependence in recruitment by age throughout the study. One recruitment chain, the longest (orange), tended to have a slightly higher proportion of those aged 25 years and over compared to other long recruitment chains (green & purple, Figure A3a), consistent with evidence of recruitment homophily by age (Table 1). The fourth longest chain (pink) tended to have a higher proportion of those aged under 25, perhaps also consistent with a lower proportion of those reporting attendance in higher education (Figure A3b). However convergence on educational attendance was achieved for the sample as a whole. Similarly, current employment converged for across the whole sample for each employment category, with no evidence of seed dependence in the proportion unemployed by recruitment chain (Figure A3c)

The cumulative weighted estimated proportion of different self-identified sexual identities did not appear to vary greatly by recruitment chain (not shown), and convergence was achieved on the overall sample albeit slightly later in recruitment than for demographic variables. Conversely there was some evidence that the cumulative proportion of transfeminine participants was still increasing as the study closed (Figure A2e), accounted for by a consistently higher proportion of transfeminine recruits in one of the larger recruitment chains active at the end of the study (not shown).

With respect to HIV outcomes, the cumulative proportion of HIV positive participants appeared to have stabilised at slightly lower weighted proportion until late in recruitment and at the end of recruitment the weighted HIV prevalence was rising slightly (Figure A2f). The proportion of those living with HIV who were virologically suppressed reached equilibrium early in the study, yet also rose slightly very late in recruitment. Whilst there was no evidence for seed dependence in bottleneck plots in cumulative HIV prevalence, convergence of one recruitment with previously low HIV prevalence did not occur until late in recruitment, and this particular recruitment chain had a systematically higher proportion of persons living with HIV who had achieved virological suppression (not shown).

#### Recruitment Homophily

Recruitment homophily (the ratio of number of recruits that have the same characteristic as their recruiter to the number we would expect if there was no homophily) was assessed for key demographic and outcome variables (Table 1). The strongest evidence of preferential recruitment of recruits with similar characteristics of the recruiter was for age and HIV status, in both cases suggesting that recruiters were more likely to identify recruits from the same age group or HIV status. With respect to age, this concurs with reported origin of most recruits from within friendship networks which can be anticipated to be assortative. The modest indication of homophily by HIV status might reflect either overlaps of identification of recruits from sexual, rather than social, networks (thus non-independent of the recruiters HIV status), or alternatively recruitment through networks formed around HIV care or prevention services.

*Table 1: Recruitment homophily estimates*

| Characteristic | Homophily estimate | $X^2$ test for independence (*p*) |
|---|---|---|
| Age group | 1.36 | $3.58 \times 10^{-10}$ |
| Educational attendance | 1.07 | 0.295 |
| Current Employment | 1.08 | 0.0724 |
| Sexual identity | 1.00 | 0.0364 |
| Gender identity | 1.00 | 0.8593 |
| HIV status | 1.16 | $6.02 \times 10^{-9}$ |
| Virological suppression | 0.92 | 0.4665 |

#### Discussion

The majority of participants reported that they knew the person who had given them a coupon, a key assumption of the RDS method (mutuality). With the exception of two seeds who recruited less than five participants, a satisfactory number of sample waves was achieved. The demographic characteristics of the achieved sample appeared to converge convincingly by the end of recruitment, however convergence of self-identified sexuality occurred later and gender identity may not have reached equilibrium within our sample size. In the absence of recruitment homophily by these factors, this suggests some seed dependence due to segregation of social and sexual networks by these factors rather than by recruitment preferences. Primary outcome estimates (HIV status and viral suppression amongst those HIV-positive) appeared to converge reasonably, although we noted minor deviations from equilibrium late in recruitment. Our observations of increases in both HIV positive status as well as proportion virologically suppressed late into recruitment, as well as modest recruitment homophily by HIV status, suggest preferential recruitment by factors related to the receipt of HIV care later in the study, such as higher age. However this may have reflected difficulties in recruitment through social activities or perceived risks in attending central Nairobi locations during civil disruption related to election disputes that occurred late in the study and close to the study site[2].

The mean age of participants was quite young: 77% of our RDS sample were <30 years of age, compared to 45% <30 years of age in the wider Nairobi population[3]. This sampling bias has been observed in other RDS surveys of MSM adults in sub-Saharan Africa[4], and is thought to reflect age and cohort effects in MSM socialising patterns as well as relative difficulties in reaching older MSM/TG in many parts of the region[5]. The sample was also slightly different to the only previous RDS study of MSM in Nairobi[6]: the 2010 sample was somewhat older (56.5% <30 years), less likely to report post-primary education than the TRANSFORM sample, yet were similarly likely to report sex work. It is unclear whether these differences represent changes in the demographics of MSM/TP populations over the interim, or reflect differences in RDS performance. Whilst diagnostics are not available for Muraguri et al), the vast majority of recruits were derived from one seed.

**NOT FOR PUBLICATION**

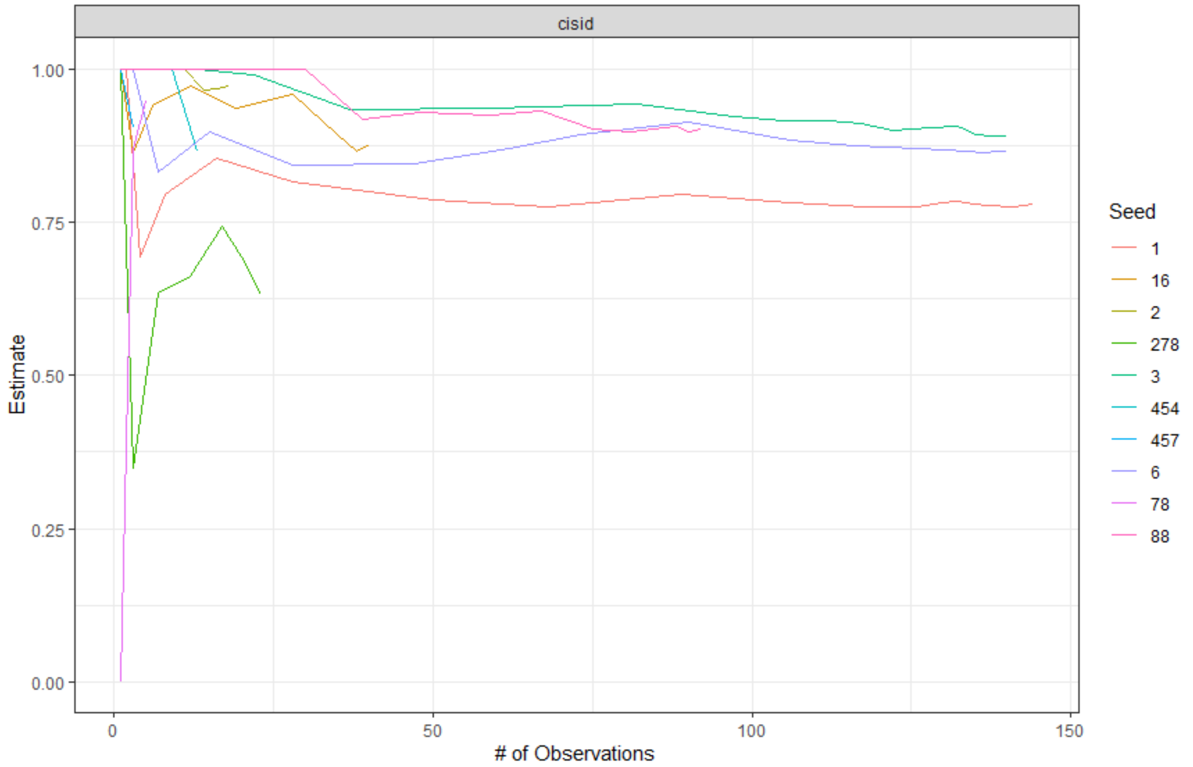


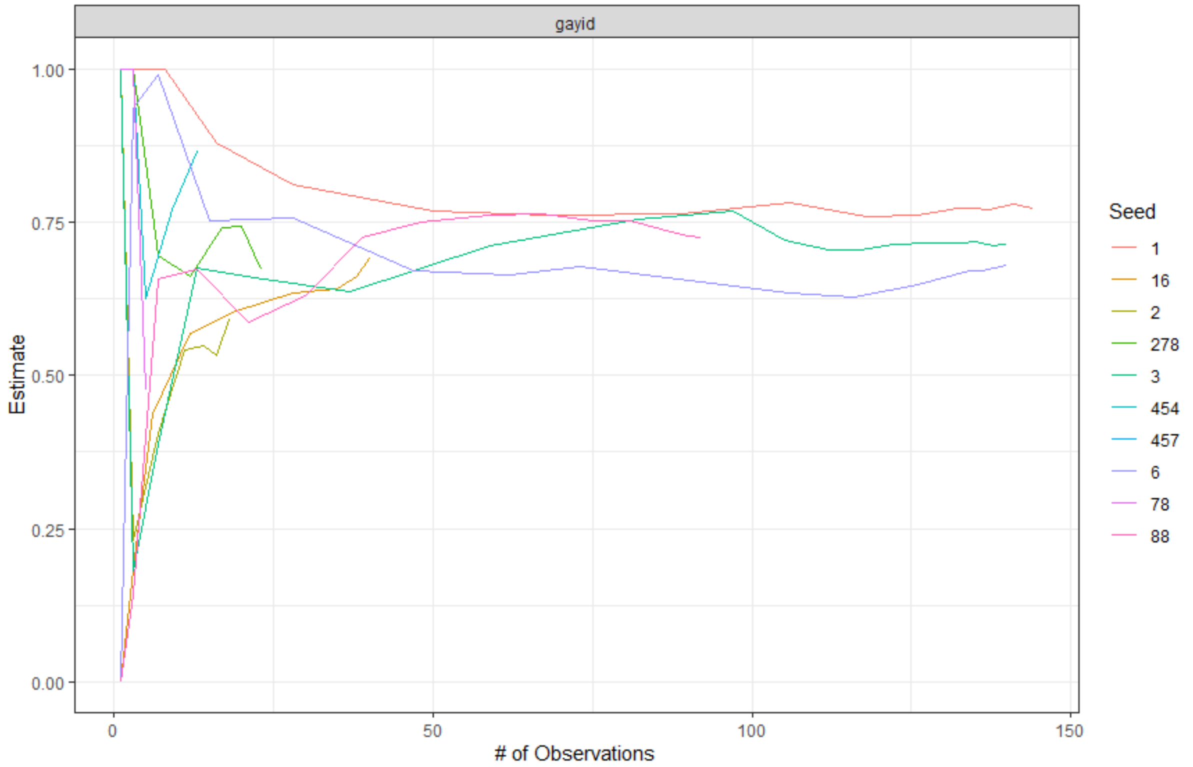

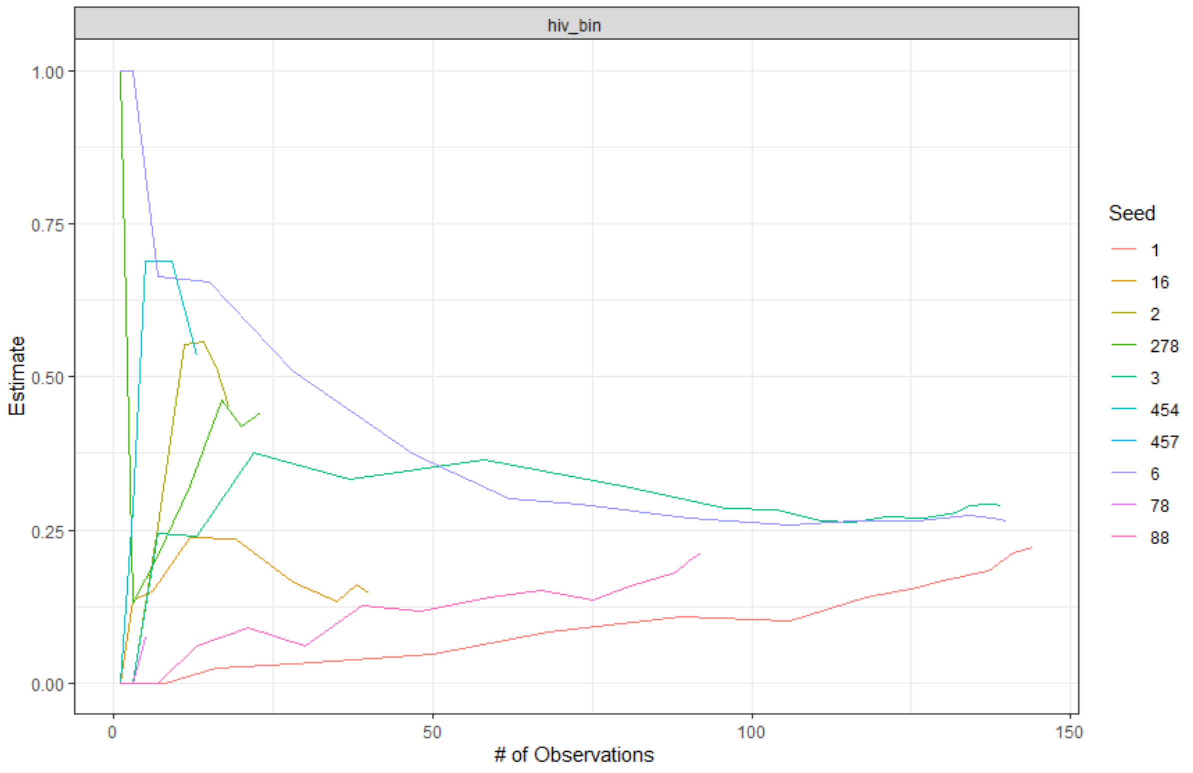
hiv_bin
Estimate
### of Observations
Seed
1
16
2
278
3
454
457
6
78
88

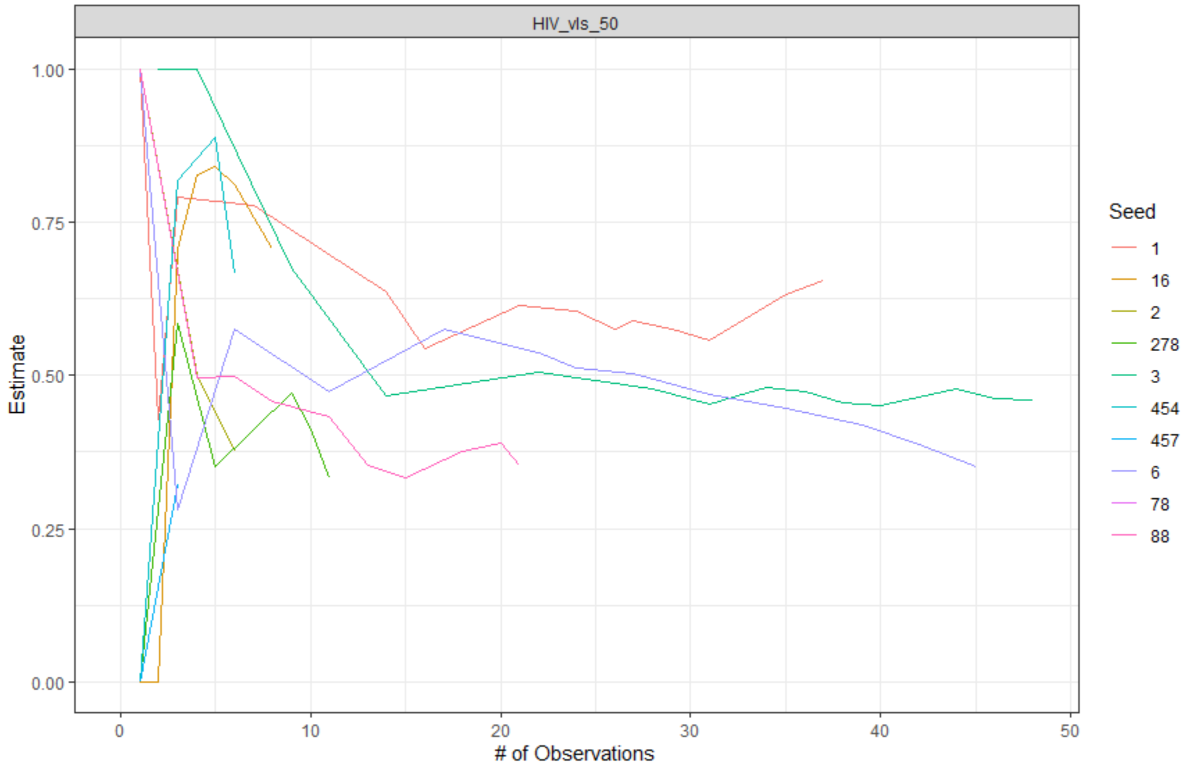
HIV_vls_50
Estimate
### of Observations
Seed
1
16
2
278
3
454
457
6
78
88
