## Appendix 2: Sensitivity analysis of source of cascade definitions and viral load cutoffs for "Age-dependent inequalities in HIV/STI burden and care receipt among men and transgender persons who have sex with men in Nairobi"

| | **Status aware** | | **Anti-retroviral therapy** | | **Viral load suppression cutoff** | | | | | |
|---|---|---|---|---|---|---|---|---|---|---|
| | Disclosed awareness of living with HIV | | Disclosed current use of ART | | <1000 copies/ml | | <200 copies/ml | | <50 copies/ml | |
| | n | %(CI) | n | %(CI) | n | %(CI) | n | %(CI) | n | %(CI) |
| **Computer assisted survey** (*self-completed*) | 137 | 69.2 (60.5-76.8) | 102 | 51.6 (43.0-60.2) | | | | | | |
| **Clinical record** (*face-to-face collected*) | 119 | 60.2 (51.5-68.4) | 115 | 57.7 (49.0-66.0) | 112 | 58.2 (49.5- 66.4) | 102 | 51.7 (43.0-60.2) | 92 | 47.4 (38.9-56.0) |
| **Composite** | 150 | 76.7 (68.3-83.3) | 129 | 65.3 (56.6-73.2) | | | | | | |
